# Protocol for a prospective feasibility study investigating phenoconversion of *CYP3A4*, *CYP2C19* and *CYP2D6* genotypes in paediatric and adolescent and young adult patients with an acute diagnosis of Hodgkin or Non-Hodgkin Lymphoma [PEGASUS]

**DOI:** 10.1101/2024.06.04.24308472

**Authors:** Rachel Conyers, Tayla Stenta, Ben Felmingham, Andrew Somogyi, Carl Kirkpatrick, Andreas Halman, Claire Moore, Dhrita Khatri, Elizabeth Williams, Roxanne Dyas, David A. Elliott, Amanda Gwee, Marliese Alexander

## Abstract

**Introduction:** Phenoconversion is the discrepancy between the predicted phenotype based on genotyping (genotype-based phenotype) and the actual phenotype influenced by non-genetic factors (clinical phenotype). Despite its potential impact on drug selection, efficacy, toxicity, and cancer treatment outcomes, research in this area is limited. This study aimed to assess the acceptability and feasibility of investigating phenoconversion using probe medications in a paediatric and adolescent and young adult oncology patient population.

**Methods and Analysis:** This prospective, single-arm, partially blinded, non-randomized feasibility study will enrol individuals aged 6-25 with a new diagnosis of Hodgkin Lymphoma or Non-Hodgkin Lymphoma. Genotyping will be performed at baseline using whole genome sequencing or targeted panel testing. Longitudinal phenotyping will be conducted throughout the cancer treatment journey using exogenous oral enzyme-specific probes, specifically subtherapeutic doses of dextromethorphan (CYP2D6) and omeprazole (CYP2C19, CYP3A4) for enzyme activity assessment. The primary outcome measure will be the proportion of patients who consent to the study and successfully complete baseline and at least two longitudinal time points with valid probe drug metabolic ratio measurements. Secondary outcomes include classification of clinical phenotypes based on probe drug metabolic ratios, probe drug safety, barriers to consent, acceptability of pharmacogenomic and phenoconversion testing, longitudinal genotype/phenotype concordance and inflammatory profiles, and investigation of patient and disease factors influencing phenoconversion.

**Ethics and Dissemination:** The ethics approval of the trial has been obtained from the Sydney Childrens Ethics Committee (2023/ETH1954). Findings will be disseminated through peer-reviewed publications and professional conference presentations.

**Trial Registration:** ClinicalTrials.gov NCT 06383338

**STRENGTHS AND LIMITATIONS OF THIS STUDY:**

- Pioneering study: This is the first study to conduct longitudinal phenotype assessments in a paediatric and adolescent and young adult oncology population.
- Primary outcome focus: The primary outcome includes patient consent and successful longitudinal probe drug derived clinical-phenotype assessments, crucial for designing future clinical trials.
- Generalisability: Conducting the study within both paediatric and adult hospital systems will enhance generalisability.
- Blinding: Probe drug metabolic ratio assessments are conducted blinded to genotype.
- Power: The trial is not powered to assess outcomes of or factors influencing phenoconversion, however secondary outcome evaluations may help prioritise outcomes/factors for further investigation.

## 1. INTRODUCTION

### 1.1 BACKGROUND AND RATIONALE

Genetic polymorphisms have been identified for several cytochromes P450 (CYP450) where their activity falls into two clearly defined populations; individuals whose rate and extent of metabolism results in poor metaboliser (PM) status and those who have faster metabolism resulting in extensive metaboliser (EM) status(1). Examples of these enzymes include CYP3A4, CYP2C19 and CYP2D6, which metabolise clinically important drugs used in acute lymphoblastic leukaemia and non-Hodgkin lymphoma therapy and supportive care. This includes drugs such as cyclophosphamide, bortezomib and omeprazole (CYP2C19); tramadol and ondansetron (CYP2D6)(2). Identifying these polymorphisms allows prediction of the clinical phenotype for drug metabolism for patients, thus presenting an opportunity to personalise patient treatment, with the ultimate goal of maximising treatment efficacy and minimising toxicity.

An individual’s variability in pharmacokinetics can be secondary to genotype, ontogeny, gender, comorbidities, inflammation and drug-drug interactions. Ontogeny is of particular importance in paediatrics whereby paediatric patients can have rapid physiological, maturate and developmental changes that alter drug metabolism and response. Most recently, there is growing recognition that there are circumstances in which there is discordance between predicted phenotype by genotyping, and actual phenotype under the influence of the aforementioned certain non-genetic factors. This mismatch has been termed ‘phenoconversion’. In this era of precision medicine where there is increasing uptake of pharmacogenomic testing, it is essential to investigate this genotype-phenotype discordance, as it may have a considerable impact on both cancer-directed and supportive care drug selection, drug efficacy and toxicity, and potentially cancer treatment outcomes for patients.

#### Phenoconversion studies in adult patients

Adult studies of non-cancer cohorts (n=27 studies), have reported that changes in the activity of CYP3A4, CYP2C19 and CYP2D6 can occur due to environmental factors, drug interactions, disease burden and inflammatory state(3). Studies of phenoconversion among patients with cancer are limited to small retrospective studies of adults with an investigatory focus on CYP2C19, but these studies have shown to report a discordance between genotype and phenotype (from genotype normal metaboliser to poor metaboliser measured by phenotype in 29% of patients) as well as differences in enzyme metabolic activity relative to non-cancer controls(1, 2). In addition, clinical scenarios that increase inflammation can result in the up-regulation of interleukin-6 (IL-6). Up-regulation of IL-6 has been shown to reduce the activity of CYP450 enzymes an exemplar of phenoconversion(3).

#### Phenoconversion studies in paediatrics

Defining CYP activity and its impact on drug response in children is challenging, particularly hampered by the difficulty in enrolling patients on prospective trials(4). Perhaps best described in the adult literature is the use of the ‘Geneva cocktail’(5) to study the phenotype across a range of CYP enzymes and transporters. This group recently published a study describing the use of CYP phenotyping and/or genotyping tests in 52 paediatric patients.

Phenotyping was performed using exogenous oral enzyme specific probes, namely, subtherapeutic doses of caffeine, bupropion, flurbiprofen, omeprazole, dextromethorphan, midazolam and fexofenadine for CYP1A2, CYP2B6, CYP2C9, CYP2C19, CYP2D6, CYP3A4/5 and P-glycoprotein activity. The study was a subpart of three total studies in adults and children(6-8). It was a retrospective study reviewing the results of patients in in-patients and out-patient settings (0-17 years) who had CYP genotyping or phenotyping performed between January 2005 and December 2020 as part of their routine care. Patients were included if phenotyping was examined using probe medications either as part of the Geneva cocktail or by administering a specific targeted probe (selective phenotyping (i.e., midazolam for testing CYP3A4 enzyme activity). Phenotype determination was based on the metabolite to parent drug metabolic ratio (MR) measured on capillary or venous samples(8).

##### Indications for Phenotype-Genotype Examination

Investigations of genotype and phenotype were most often carried out because of (i) insufficient response to treatment; (ii) low plasma concentration; or (iii) an adverse drug reaction. The majority of prescribed medications for which phenotype/genotype examination was sought were for immunosuppressants, antipsychotics, analgesics, antiepileptics, antithrombotics and antifungals(8).

##### Phenotype-Genotype Concordance Rates

In this combined retrospective study, 52 patients (mean age 9.7 (+/- 5.4 years, range 1 month- 17 years) underwent genotype or phenotype testing over a period of 2 decades. Of the 52 patients, 42% underwent simultaneous genotyping and phenotyping, 23% underwent phenotyping only and 35% underwent genotyping only. In total 17 patients were able to be analysed for combined genotype and genotype (mean age 9.3 +/- 5.4 years). The concordance rates between phenotype and genotype were highly variable ranging from 80% for CYP2C9 to 33% for CYP3A5. Interestingly, in ∼50% of the non-concordant groups, the phenoconversion could be explained by a drug-gene interaction (drug-induced phenoconversion). Furthermore, in all the CYP2D6 examined patients, the phenotype exhibited increased activity in comparison to the genotype, raising the question of other potential causes. The results of the study showed that the patients genotype:phenotype mismatch explained the clinical reason for phenotype testing (i.e., low therapeutic drug monitoring levels) in 60% of cases.

##### Probe Drug Administration and Reported Adverse Events

Most patients within the above study received targeted phenotyping (rather than the full Geneva cocktail) focusing on up to 3 CYP enzymes. Twelve patients received the full Geneva cocktail. The most frequently examined enzymes were CYP3A4 (n=27) followed by CYP2D6 (n=23). Of note, no adverse events were reported following targeted or Geneva Cocktail administration(8).

##### Rationale for the study of phenoconversion in paediatrics and paediatric oncology

The paediatric population is of particular relevance for phenoconversion in the paediatric oncology setting with ontogeny, drug-drug interactions and the potential for inflammatory events either secondary to disease or infection. Furthermore, there is wide inter-individual variability. These factors combined make safe but also effective drug cancer and supportive care medication delivery particularly challenging. At any point in time during cancer therapy, the milieu of factors could lead to increased clearance of a medication and inefficacy, or conversely, decreased clearance and drug toxicity. Determining and characterising their influences on drug metabolising enzyme activity to guide therapy is of particular importance in the era of personalised medicine. This is particularly relevant in paediatric oncology where (i) there are some high-risk diseases with unexplained poor response to molecular- or chemo-therapy and (ii) where survivorship and long-term therapy related toxicities is of particular concern.

##### Rationale for the study of CYP3A4, CYP2C19 and CYP2D6 enzymes

The decision to assess CYP3A4, CYP2C19 and CYP2D6 enzymes was based both on critical roles in the metabolism of many oncology and supportive care medicines(9, 10) and the availability of accessible, safe, and easily prepared, administered, and measured probe drugs(11)

##### Rationale for inclusion of microsampling

Longitudinal phenotype assessments will be conducted using traditional blood draw methods (central venous access device or peripheral blood) and Volumetric Absorptive Microsampling (VAMS). The inclusion of VAMS aims to demonstrate its potential for patient self-testing, thereby minimizing the need for invasive testing and reducing required hospital/pathology visits. This approach is intended to improve the acceptability of future clinical trials from the patient perspective, by offering a less burdensome and more convenient option for ongoing phenotype monitoring. VAMS was selected over traditional dried blood spot (DBS) microsampling for collection of fixed blood volumes and superior accuracy and reliability(12).

### 1.2 OBJECTIVES

#### 1.2.1 Hypothesis

Probe medications can be administered to measure phenoconversion at multiple timepoints in the cancer journey in a paediatric/AYA patient population.

#### 1.2.2 Primary Objective

To test acceptability and feasibility to further studies of phenoconversion using probe medications in a paediatric/AYA oncology patient population.

#### 1.2.3 Secondary Objective

1. Classify CYP phenotypes according to metabolic ratio (MR) of probe drug administration.
2. To understand barriers to consent and acceptability of the study, from patient and clinician perspectives.
3. To understand barriers to viable sample collection, shipping, processing, and timely reporting.
4. To understand safety of phenoconversion testing using probe drug administration.
5. Describe phenoconversion in terms of genotype and phenotype mismatch across longitudinal timepoints.

## 2.0 TRIAL DESIGN

**METHODS: PARTICIPANTS, INTERVENTIONS, OUTCOMES**

### 2.1 Trial Design

Prospective single arm partially blinded and non-randomised feasibility study.

The study will be conducted in patients (6-25 years of age) with Hodgkin lymphoma or non-Hodgkin lymphoma as exemplar cohort, but with the understanding that cancer directed and supportive care medicines of the CYP3A4, CYP2C19, and CYP2D6 metabolic pathways, are commonly utilised for the treatment of many paediatric, adolescent, young adult, and adult cancers. The acceptability and feasibility of this study will inform future studies in phenoconversion within the cancer population to direct more personalised precision medicine.

### 2.2 Sponsorship, Trial Registration and Version

The trial is sponsored by the Murdoch Children’s Research Institute and registered with clinicaltrials.gov NCT 0638338 [first registered 21.03.2024]. It is approved Sydney Childrens Ethics Committee (2023/ETH1954). The study is currently version 2 26.04.2024.

## 3.0 STUDY SETTING

PEGASUS will be conducted at two tertiary institute in Victoria, Australia. The study is focussed on enrolling patients within oncology units within a paediatric unit [The Royal Children’s Hospital, Melbourne] and an adult unit [Peter MacCallum Cancer Institute].

## 4.0 ELIGIBILITY CRITERIA

### 4.1 Participants inclusion criteria

- Age 6-25 years of age.
- New diagnosis of Hodgkin Lymphoma or Non-Hodgkin Lymphoma.
- Able to swallow and absorb oral or nasogastric tube (NGT) administration of probe drugs.
- Able to provide written informed consent.

### 4.2 Participants exclusion criteria

- Failure to comply with inclusion criteria.
- Has a known previous allergy to any of the probe medications (i.e., omeprazole or dextromethorphan).
- Common Terminology Criteria for Adverse Events (CTCAE) version 5.0 Grade IV end organ dysfunction (i.e., hepatic, renal, gastrointestinal).
- Had previous oncological treatment (not first cancer diagnosis).
- Is a clinically unstable patient requiring incentisive care admission in high-risk circumstances will not be considered eligible for consent.
- Any patient requiring urgent initiation of anti-cancer treatment outside hours where a member of the study staff is unable to approach the parent/guardian or participant for consent prior to commencing anti-cancer therapy will be ineligible for consent.
- Unable to provide written informed consent.

## 5.0 INTERVENTIONS

### 5.1 Treatment arms

This is a single arm study where all participants will undergo genotyping of their CYP3A4, CYP2C19 and CYP2D6 enzymes either via whole genome sequencing or targeted panel testing. All participants are given the intervention (two sub-therapeutic probe drugs) with subsequent blood sampling (2 hours later) to test enzyme activity (phenotyping).

PEGASUS timepoints are as follows:

**(i)** Prior to commencing first lymphoma chemotherapy cycle
**(ii)** 2 months (week 8)
**(iii)** 4 months (week 16)
**(iv)** Completion of therapy (> week 16)
**(v)** Up to a maximum of 2 febrile neutropenic episodes.

A minimum of 24 hours must occur between study visits.

## 6.0 OUTCOMES

Objective and outcome measures are detailed in Table 1.

**Table 1:** Trial Objectives and Outcomes. Abbreviations: AE: Adverse Event; AYA: Adolescent and young adult; CRP: Cytokine Reactive Protein; ESR: Erythrocyte Sedimentation Rate; LDH: Lactate Dehydrogenase; MR: Metabolic ratio; PCT: Procalcitonin; TFA: Theoretical Framework of Acceptability; VAMS: Volumetric absorptive microsampling;

| OBJECTIVE | OUTCOME & OUTCOME MEASURE |
| --- | --- |
| **Primary** |  |
| To test acceptability and feasibility of further studies in phenoconversion using probe medications in a paediatric/AYA oncology patient population. | To determine the proportion of patients who consent to study and complete baseline and at least two longitudinal timepoints with successful measurement of probe drug MR. |
| **Secondary** |  |
| To classify CYP phenotypes according to MR of probe drug administration. | 1. Proportion of patients completing all longitudinal blood sampling.<br>2. Proportion of patients with successful detection of probe drug overall and at each sampling timepoint.<br>3. Proportion of patients where phenotype can be classified according to MR overall at each sampling timepoint.<br>4. Proportion of patients using VAMS technique where phenotype can be classified according to MR overall and at each sampling timepoint in comparison to blood draw. |
| To apply the TFA to understand barriers to consent and acceptability of participating in pharmacogenomic and phenoconversion testing from patient and clinician perspectives | 1. Reported acceptability of the study measured via survey at study completion from patient, clinician and clinical trial coordinator perspective.<br>2. Proportion of patients declining consent |
|  | and classification of these according to pre-defined parameters at time of consent.<br>**3.** Determine the proportion of patients declining consent or withdrawing consent and classification of these according to pre-defined parameters at time of consent. |
| To understand safety of phenoconversion testing using probe drug administration. | Percentage of patients experiencing an AE during probe drug administration. |
| To describe phenoconversion in terms of genotype and phenotype mismatch across longitudinal timepoints. | Incidence of genotype and phenotype mismatch overall across longitudinal timepoints. |
| To document patient and disease factors that may influence phenoconversion. | Proportion of patients with documentation of all parameters:<br>(i) Disease staging and biomarkers of extent of disease (ESR, LDH).<br>(ii) Systemic inflammatory state (CRP, procalcitonin and cytokine panel).<br>(iii) Current medications involving the CYP P450 pathway.<br>(iv) Demographics (age, gender, ethnic background).<br>(v) Other environmental factors (alcohol, smoking, vitamin D status). |
| To describe the longitudinal inflammatory profile of patients with Hodgkin or non-Hodgkin Lymphoma. | Inflammatory profile measured by CRP, PCT and cytokine panel. |

### 6.3 Methodology for Primary Outcome Phenotyping

#### 6.3.1 Using Probe Medications for Phenotyping

Phenotyping will be performed using exogenous oral enzyme-specific probes; namely subtherapeutic doses of dextromethorphan (CYP2D6), omeprazole (CYP2C19) (CYP3A4) for enzyme activity assessment.

Several phenotype cocktails have been developed using probe drugs to study human CYP P450 enzymes. One well known cocktail that has been used is known as the Genevea cocktail consisting of caffeine, bupropion, flurbiprofen, dextromethorphan, midazolam, omeprazole, and fexofenadine). The dose of omeprazole and dextromethorphan used in this study was 10 mg of each drug (below the daily therapeutic dose).

#### 6.3.1 Omeprazole

A proton pump inhibitor with therapeutic effect achieved by binding to the hydrogen/potassium ATPase enzyme system (proton pump), inhibiting both stimulated and basal acid secretion. It has several indications and is often used to treat peptic ulcer disease, gastroesophageal reflux disease, and routinely in oncology patients for the prevention and treatment of upper gastrointestinal adverse events. Proton pump inhibitors are generally well tolerated. Further information is available in Supplemental File 1.

#### 6.3.2 Potential omeprazole adverse effects

**Table 2:** Potential adverse effects according to the Australian Medicines Handbook, July 2023.

| Common (> 1%) | Infrequent (0.1 – 1%) | Rare (< 0.1%) |
| --- | --- | --- |
| headache, nausea, vomiting, diarrhoea, abdominal pain, constipation, flatulence. | rash, itch, dizziness, fatigue, drowsiness, insomnia, dry mouth. | gynaecomastia, myalgia/arthralgia, acute interstitial nephritis, peripheral oedema, hepatitis, jaundice, thrombocytopenia, leucopenia, hypersensitivity reactions (i.e., anaphylaxis, Stevens-Johnson syndrome), subacute cutaneous lupus |
|  |  | erythematosis (very rare). |

#### 6.3.3 Dextromethorphan

An opioid derivative (but lacking any opioid activity) dextromethorphan is known to be an *N-* methyl-d-aspartate (NMDA) <u>receptor antagonist</u>. However, dextromethorphan binding sites are not limited to the known distribution of NMDA receptors. Dextromethorphan also inhibits the reuptake of serotonin. The use of dextromethorphan is not recommended by the Australian Medical Association for use in children under 6 years of age. Further information is available in Supplemental File 2.

#### 6.3.4 Potential dextromethorphan adverse effects

**Table 3:** Potential adverse effects according to the Australian Medicines Handbook, July 2023.

| Common (> 1%) | Infrequent (< 0.1%) | Rare |
| --- | --- | --- |
| drowsiness, constipation,<br>nausea, vomiting. | respiratory depression. | This box is intentionally<br>blank. |

### 6.4 Dosage and route of administration

Probe drugs will be administered orally or via nasogastric tube. Both probe drugs will be administered at the same time, one drug after the other, any order. All patients will receive the probe drug dose which is below the safest sub-therapeutic dose for the lowest age range (6-year-old). To ensure the safest subtherapeutic dose is administered to individuals that are not routinely taking a dose of the probe drug, a dose of 5 mg omeprazole and 5mg dextromethorphan will be administered to all participants (Table 4). This is half the dose used in the Geneva cocktail where MR have been successfully measured, and the dose has been safe and well tolerated with low incidence of adverse events from a single oral dose.

**Table 4:** Probe Drug dosing and administration.

|  |  |  |  |
| --- | --- | --- | --- |
|  | **PEGASUS**<br>**subtherapeutic dosing** | **Australian Medical**<br>**Handbook**<br>**omeprazole** | **Standard**<br>**therapeutic dose**<br>**based on 21kg** |

**Table 4: Probe Drug dosing and administration**
|  |  | therapeutic dosing |  |
| --- | --- | --- | --- |
| Omeprazole | 5mg once (10mg tablet dispersed in 10mL water/juice with 5mL administered). | Birth(at term)<br>(0.7 mg – 1.4 mg/kg DAILY) *<br>All patients<br>5 mg daily to 20 mg TWICE daily. | 10mg – 20mg once daily |
| Dextromethorphan | 5mg once (2.5mL liquid). | 6 yrs to adult *<br>5mL – 20mL DAILY<br><u>Concentrations may vary if BISOLVON<sup>®</sup> brand is not used.</u> | 5mL up to FOUR times daily. |

\* The average weight of a 6-year-old patient according to the Australian Medical Handbook is 21kg.

Probe drugs will be dispensed by site designated pharmacy departments in accordance with local pharmacy standard operating procedures (SOPs) for dispensing of study medication. Omeprazole can be dispensed as any commercially available form as approved for use in Australia by the Therapeutic Goods Administration throughout the study trial. Individual doses should be dispensed in an amber glass bottle. Dextromethorphan hydrobromide should be administered using the BISOLVON© product as this is the only single agent commercially available product in Australia approved by the Therapeutic Goods Administration. If this specific brand is unable to be obtained by the pharmacy department a suitable combination product must be approved by the study PI. Individual doses should be dispensed in a single oral syringe for administration.

#### 6.4.1 Cases where a participant is already taking a probe drug routinely

Omeprazole is often given as a regular medication for patients receiving chemotherapy, particularly those taking steroid medication (i.e., prednisolone, dexamethasone). In instances where a patient is already taking a probe medication at therapeutic doses (most likely omeprazole rather than dextromethorphan) there will be no need to administer a second sub-therapeutic drug probe dose. The enzyme activity can be measured regardless of the administered dose using the MR approach. The different dosing regime will be recorded in study case report forms.

#### 6.4.2 Minimum documentation of medication information by pharmacy

For each probe drug the brand name, batch and expiry must be recorded by the dispensing pharmacy site for study reference if required. This information should be documented as part of good routine medication dispensation. All dispensing of medication supply will be verified by a second staff member. The probe drugs will be dispensed in accordance with the investigator’s prescription. Pharmacy will be responsible for notifying the study PI if there are any concerns or issues relating to the safe handling and storage of the studies probe drugs (i.e., temperature excursions, damaged packaging).

### 6.5 Drug Levels

#### 6.5.1 Measuring activity of CYP2C19 and CYP3A4 by measuring the MR of omeprazole

The MR of 3 CYPS (3A4, 2C19, 2D6) will be measured 2 hours (+/-30 minutes) following probe drug administration. MR will be measured by blood draw from CVAD or peripheral blood draw and VAMS. Blood draw will be taken in a standard EDTA tube and both VAMS and EDTA will be stored at room temperature until ready for processing.

CYP2C19 metabolises R-omeprazole to the hydroxy metabolism which represents almost 90% of R-omeprazole metabolism. CYP2C19 metabolises S-omeprazole to the 5-O-desmethylmetabolite representing 40% of overall S-omeprazole metabolism.

CYP3A4 metabolises S-omeprazole to the sulphone metabolite representing ∼ 57% of overall S-omeprazole metabolism. Therefore, administration of omeprazole and measurement of hydroxy omeprazole will allow approximation of CYP2C19 phenotype, and the sulphone metabolite will give CYP3A4 phenotype.

#### 6.5.2 Measuring activity of CYP2D6 by measuring the MR of dextromethorphan

Dextromethorphan and its primary metabolites (CYP2D6-mediated dextrorphan; CYP3A4-mediated 3-methoxymorphinan) and secondary metabolite 3-hydroxymorphinan will be quantified in plasma using a modification of the original method from the Somogyi (CI) laboratory which originally used fluorescence detection (PMID 2305428) and which was easily able to identify CYP2D6 PMs (PMID 8841152) as the MR was over 1000-fold different between EMs and PMs. The primary modification being use of mass spectrometry instead of fluorescence, which will enhance sensitivity.

#### 6.5.3 Determining Phenotype of CYP2D6, CYP2C19 and CYP3A4 from the MR

Phenotypic classification is based on the metabolite to parent drug metabolic ratio (MR). This is defined as the concentration of the metabolite divided by the concentration of the substrate according to a validated method using liquid chromatography tandem mass spectrometry quantification. The MR can then be used to give a representative phenotype (i.e., poor metaboliser (PM), intermediate metaboliser (IM), normal metaboliser (NM) or ultra-rapid metaboliser (UM).

Threshold levels for phenotype assessment have been detailed previously by Lenoir et al(13) (See Table 5). For noting in the Lenoir et al study CYP3A4 reference ranges for MR and SD were determined using midazolam as the probe medication.

**Table 5:** Threshold levels for phenotype assessments.

| | **PM**<br>(Mean MR ± SD) | **IM**<br>(Mean MR ± SD) | **NM**<br>(Mean MR ± SD) | **UM**<br>(Mean MR ± SD) |
| --- | --- | --- | --- | --- |
| CYP2C19 | 0.18 ± 0.10 | NA | 0.76 ± 0.46 | 5.42 ± 2.46 |
| CYP2D6 | 0.05 ± 0.02 | 0.46 ± 0.41 | 2.41 ± 1.79 | NA |
| CYP3A4 | 0.22 ± 0.07 | NA | 0.57 ± 0.25 | 3.74 ± 1.50 |

Abbreviations: NA = Not applicable; MR = metabolic ratio; SD = standard deviation.

### 6.6 Methodology for Genotyping

All participants will have genotyping performed for pharmaco-genes CYP3A4, CYP2C19 and CYP2D6 via either whole genome sequencing [WGS] (if patient has had previous WGS or gene panel testing undertaken prior to PEGASUS enrolment) or genotyping via a targeted gene panel looking at CYP3A4, CYP2C19 and CYP2D6 for patients who have not already gene sequencing results available. This will involve either a blood sample being taken or buccal swab. Some participants may have already consented to the MARVEL-PIC (HREC 89083) or PRECISION-ITS (PMCC HREC 101174) studies and thus will have the genotyping result already available and reported within their EMR. These are existing collaborations between A/Prof Conyers and Dr Marliese Alexander. Bloods will not be re-taken if this result is already available, and the results already taken will be utilised for this study.

## 7.0 PARTICIPANT TIMELINE

The study schema (Figure 1) and schedule of assessments (Table 6) provide an overview of study visits and procedures.

**Figure 1:**
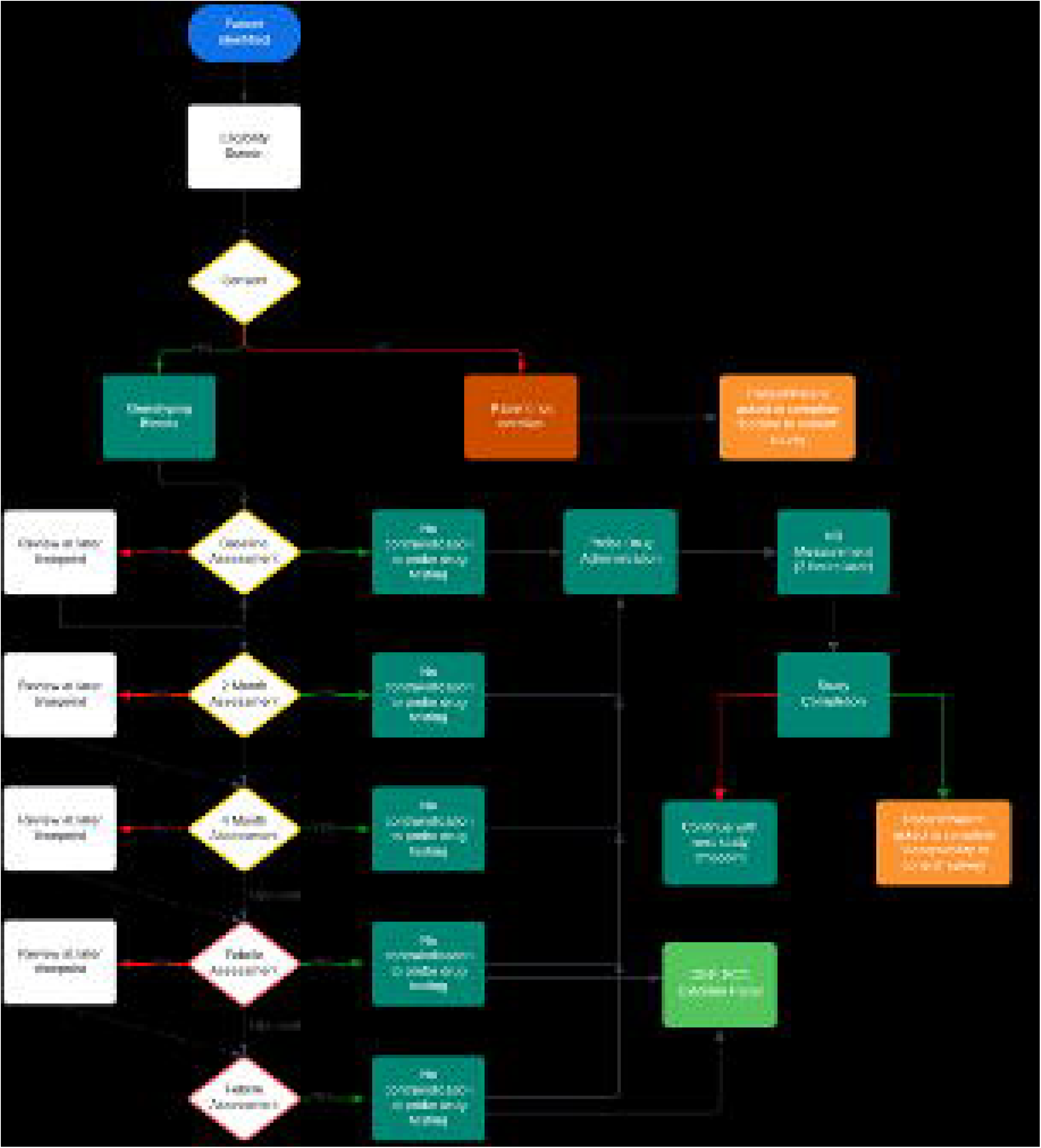
Schema of PEGASUS Study Overview and Procedures. Contraindication to probe drug testing is assessed in consultation with paediatric oncologist in collaboration with treating pharmacist. If patient misses a timepoint they automatically move to the next timepoint in the study. Probe drug administration includes administration of dextromethorphan and omeprazole as described in section 6.3.1 and 6.3.2 Phenotyping (MR) measurements are performed 2 hours after probe drug administration. Febrile episodes up to a maximum of two episodes will be included in the study.

**Table 6:** Overview of Study Visits and Procedures.

| PEGASUS | TRIAL PERIOD |  |  |  |  |  |  |
| --- | --- | --- | --- | --- | --- | --- | --- |
|  | Enrolment | Timepoints |  |  |  |  | Close-out |
| TIME POINT** | $-t_1$ | $t_1$ | $t_2$ | $t_3$ | $t_4$ | $T_{5/6}$ | $T_7$ |
| **ENROLMENT:** |  |  |  |  |  |  |  |
| **Eligibility screen** | X |  |  |  |  |  |  |
| **Informed consent** | X |  |  |  |  |  |  |
| **Weight/height/vital signs** | X |  |  |  |  |  |  |
| **Medical history, Clinical Examination and EMR review** |  | X(a) | X(a) | X(a) | X(b) | X(b)<br>) | X(a) |
| **INTERVENTIONS :** |  |  |  |  |  |  |  |
| **Blood test: genotyping** | X (C) |  |  |  |  |  |  |
| **Sub-therapeutic dosing omeprazole & dextromethorphan** |  | X(d) | X(d) | X(d) | X(d) | X(d)<br>) | X(d) |
| **Blood test: Phenotyping** |  | X(e) | X(e) | X(e) | X(e) | X(e)<br>) | X(e) |
| **Blood test: CRP, PCT and cytokine panel** |  | X(f) |  |  |  | X(f) | X(f) |
| **Acceptability Survey (TFA)** |  |  |  |  |  |  | X |

**Timepoints:** $t_1$ prior to enrolment, $t_2$ at diagnosis, $t_3$ at 2 months, $t_4$ at 4 months, $t_{5/6}$ at maximum of two febrile episodes within a 4-month period, $t_7$ at study conclusion.
- a. Participant admitted to day oncology space, or if already an inpatient will be seen on the ward; will then be administered dosing of dextromethorphan/omeprazole. Visit will take 2.5 hours. - b. Participant will be admitted to ward or day oncology fast track unit with febrile episode. Will then be administered dosing of dextromethorphan/omeprazole. visit will take 2.5 hours. - c. If a participant is already enrolled on the “MARVEL-PIC” or “PRECISION” protocol, this test will be omitted as results will already be available. - d. Medication will be dispensed as outlined in section 5.2 following medication reconciliation. - e. Bloods to measure enzyme activity following probe drug administration to be taken by central venous access device and VAMS in consultation with the nursing team. - f. Inflammatory profile to be done on a maximum of two febrile admissions during the study period, together with $t_2$ and $t_7$ .

## 8.0 SAMPLE SIZE

As the primary outcome is feasibility and acceptability a sample size estimate is not required, however it should be noted that the investigators estimate that the recruitment period will be either adequate to permit an assessment of feasibility and acceptability, or to determine that a trial is not feasible due to low enrolment.

## 9.0 RECRUITMENT

### 9.1 Achieving Sample Size

This study is aiming to enrol 10 patients. To mitigate the risk of poor enrolment we will align recruitment with routine hospital care and admissions to minimize burden to the patient.

Understanding acceptability of the trial is an important endpoint and, as such, we will try and understand failure to recruit using short pre-scripted questions at the time of consent.

### 9.2 Recruitment

Clinical trial assistants or academic pharmacists at each centre will screen eligible patients through the outpatient or inpatient department. The duration of the recruitment period is estimated to be a 24-month interval depending on each centre’s recruiting rate. No financial incentives will be provided to trial investigators or patients for enrolment in the recruitment period.

## 10. ALLOCATION

### 10.1 Sequence generation and Concealment

This study is partially blinded and non randomised.

### 10.2 Implementation

Following consent, the research assistant or academic pharmacist enrolling the patient will enter details onto the study database.

### 10.3 Blinding

The patients, pharmacists and physicians are unblinded to the pharmacogenomic results. The scientific team assessing probe medication MR will be blinded to genotype.

## METHODS: DATA COLLECTION, MANAGEMENT, ANALYSIS

### 11.0 DATA COLLECTION PRIMARY OUTCOME AND SECONDARY OUTCOMES

#### 11.1 Eligibility Screen

The screening visit will take place once a new Hodgkin Lymphoma or Non-Hodgkin Lymphoma diagnosis is made in an eligible patient.

Prior to any study specific procedures being performed, written informed consent will be obtained from the patient/guardian(s). The screening visit will take place over 1 day or a number of days depending on the patient/family schedule. The following assessment will be undertaken at the screening visit.

- Inclusion/exclusion criteria check
- Written informed consent
- Medical History including medication reconciliation.
- Weight/height/vital signs
- Review of EMR
- Blood draw or buccal swab for genotyping of pharmacogenes (2mL – 5mL) in an EDTA tube if WGS or panel PGx data not already available for patient on MARVEL-PIC [National Randomised Controlled Trial PGx Implementation – HREC 89083] or PRECISION [Peter MacCallum PGx study – HREC 101174] studies. Absolute minimum required is 2mL for WGS.

**<u>NOTE:</u>** In the unlikely event that the PI of the study is on service at the RCH when there is a potential eligible patient, the PI will ask another liquid stream team member of the Children’s Cancer Centre to take over care of the patient to avoid any conflict of interest. At the PIs study site all Hodgkin’s patients are treated by the solid stream medical team leaving only non-Hodgkin’s patients to potentially fall under the PIs primary care. This would happen prior to the patient/guardian(s) being approached for study consent.

**Timing of Assessment:** t_-1_ at enrolment.

**Administration of Assessment:** PI or Academic pharmacist and bloods taken by nursing or pathology staff (for peripheral or CVAD blood samples) and performed by trained Academic Pharmacist or RA (for VAMS samples).

#### 11.2 Phenotyping visits [Standard]

Visit to be conducted by PI, delegated AI, or Academic pharmacist.

These include the following timepoints: t_1_ prior to enrolment, t_2_ at diagnosis, t_3_ at 2 months, t_4_ at 4 months and at t_7_ at study conclusion.

Prior to a phenotyping assessment the following assessments will be undertaken:

- Vital signs/weight/height
- Review of medical history
- Clinical Examination
- Medication Reconciliation
- Review of EMR

Following these assessments, the participant will be deemed suitable for probe drug administration and two hours later bloods for phenotyping via (i) central venous access device and (ii) VAMS.

##### Peripheral or Central Venous Access Device bloods

Enzyme activity will be determined by taking 2mL – 5mL of blood (EDTA) 2 hours after administration of the probe drug. The patients unique study identifier will be attached to the blood tube along with the sampling timepoint. These will also be annotated within the *REDCap* database.

Immediately after the bloods are taken samples will be centrifuged at 2000g and samples will be transferred to polypropylene microtubes and stored at -20 degrees Celsius until MR analysis.

##### Volumetric Absorptive Microsampling (VAMS)

This approach uses minute amounts of dried blood obtained via VAMS prior to LC-MS analysis. A VAMS device is used to sample 20μl of blood. The VAMS uses an innovative microsampling device, constituted by a hard plastic handle fitted with a detachable tip made of proprietary, porous polymeric material(14). The tip is capable of absorbing a field liquid sample volume according to its size and independently of sample density or viscosity. After absorption the specimen is left to dry and is then ready for storage(14). The patients unique study identifier will be attached to the VAMS along with the sampling timepoint. These will be annotated in the *REDCap* database. All samples will be stored at room temperature prior to batch processing for MR analysis.

###### Timing of Assessment after probe drug

Blood should be drawn as close as possible to 2 hours after probe administration with an acceptable window of +/-30minutes. If for any reason blood draw does not occur in this window (1.5 to 2.5 hours after probe administration), blood draw should still be attempted up to 6 hours, with documented protocol deviation as delayed assessment. No blood draw should be attempted beyond 6 hours after probe drug administration, with documented protocol deviation as missed assessment. For all blood draws accurate documentation of the time of probe drug administration and blood draw is imperative.

###### Timing of Assessment after enrollment

t_2_ at diagnosis, t_3_ at 2 months, t_4_ at 4 months, t_5/6_ at a maximum of two febrile episodes within 4-month period and at t_7_ at study conclusion (when finished treatment).

###### Administration of Assessment

PI or Academic pharmacist and bloods taken by nursing or pathology staff (for peripheral or CVAD blood samples) and performed by trained Academic Pharmacist or RA (for VAMS samples).

#### 11.3 Phenotyping visits [Febrile Illness]

These study visits will occur without scheduling t_5/6_ at maximum of two inpatient febrile episodes within 4-month period of enrolment. The PI of the study, academic pharmacists, and research assistants as part of the research team will be notified of enrolled patients being admitted with febrile illnesses by collaborating physicians on the ward or the nurses within oncology fast track clinic.

Prior to a phenotyping assessment the following assessments will be undertaken:

- Vital signs/weight/height
- Review of medical history
- Clinical Examination
- Medication Reconciliation
- Review of EMR

Following these assessments by a paediatric or adult oncologist and academic pharmacist within the team, a check will be undertaken with the treating medical team to gain consent for proceeding with dosing of probe medications. Once this is confirmed probe drug administration will occur and two hours later bloods for phenotyping (2mL – 5mL) via central venous access device.

##### Timing of Assessment after probe drug

Blood should be drawn as close as possible to 2 hours after probe administration with an acceptable window of +/-30minutes. If for any reason blood draw does not occur in this window (1.5 to 2.5 hours after probe administration), blood draw should still be attempted up to 6 hours, with documented protocol deviation as delayed assessment. No blood draw should be attempted beyond 6 hours after probe drug administration, with documented protocol deviation as missed assessment. For all blood draws accurate documentation of the time of probe drug administration and blood draw is imperative.

##### Timing of Assessment after enrollment

t_5/6_ at a maximum of two febrile episodes within a 4-month period.

##### Administration of Assessment

PI or Academic pharmacist and bloods taken by nursing or pathology staff (for peripheral or CVAD blood samples) and performed by trained Academic Pharmacist or RA (for VAMS samples).

#### 11.4 Description of Procedures

##### 11.4.1 Genotyping

All participants will have genotyping performed for pharmaco-genes CYP3A4, CYP2C19 and CYP2D6 via either WGS (if patient has had previous WGS or gene panel testing undertaken prior to PEGASUS enrolment) or genotyping via a targeted gene panel looking at CYP3A4, CYP2C19 and CYP2D6 for patients who have not already gene sequencing results available. This will involve either a blood sample being taken or buccal swab. Some participants may have already consented to the MARVEL-PIC (HREC 89083) or PRECISION (PMCC HREC 101174) studies and thus will have the genotyping result already available and reported within their EMR these are existing collaborations between A/Prof Conyers and Dr Marliese Alexander. Bloods will not be re-taken if this result is already available, and the results already taken will be utilised for this study.

Clinical Pharmacogenetics Implementation Consortium (CPIC) and Dutch Pharmacogenetics Working Group (DPWG) guidelines will be used to predict and classify the participant’s phenotype based on individual genotype when this result is available.

**Timing of Assessment:** This is performed at enrolment (t_-1_).

**Administration of Testing:** Testing ordered by PI/Academic pharmacist in the EMR and bloods taken by nursing or pathology staff (for peripheral or CVAD blood samples) and performed by trained Academic Pharmacist or RA (for VAMS samples).

##### 11.4.2 Probe Drug Administration and bloods for phenotyping

Participants will receive probe drug administration as outlined in section 6.3.1. Two hours (+/- 30minutes) following probe drug administration bloods will be drawn from the participants central venous access device or via VAMS for phenotyping of CYP2C19, CYP3A4 and CYP2D6.

Volumetric absorptive micro sampling (VAMS) is a recently developed sample collection method that enables single-drop blood collection in a minimally invasive manner. Blood biomolecules can be extracted and processed for analysis via mass spectrometry.

**Timing of Assessment:** t_2_ at diagnosis, t_3_ at 2 months, t_4_ at 4 months, t_5/6_ at a maximum of two febrile episodes within a 4-month period and at t_7_ at study conclusion (when finished treatment).

**Administration of Testing:** Testing ordered by PI/Academic pharmacist and bloods taken by nursing or pathology staff (for peripheral or CVAD blood samples) and performed by trained Academic Pharmacist or RA (for VAMS samples).

#### 114.3 Inflammatory profiling

Serum levels of PCT, CRP and cytokine analysis will be performed at baseline and at a maximum of 2 febrile episodes within a 4-month period. Concentrations of CRP will be measured from lithium heparin whole blood sample, directly after blood collection using latex enhanced immunoturbidimetry. Blood samples will be centrifuged at 2,000 g and 4°C for 10 minutes and serum samples stored at –80°C until analysis. A panel of 48 inflammatory cytokines will be measured using the Bio-RAD Bio-Plex Pro Human cytokine screening panel (bio-rad.com/en-au). This screening panel detects a comprehensive set of studied and biologically relevant cytokines including those shown to be predictive of severe infection in children with cancer and febrile neutropenia(15). This technique allows simultaneous analysis of multiple samples together using small sample volumes.

**Timing of Assessment:** t_2_ at diagnosis and t_5/6_ at a maximum of two febrile episodes within 4-month period and at t_7_ at study conclusion.

**Administration of Testing:** Testing ordered by PI/Academic pharmacist and bloods taken by nursing or pathology staff.

##### 11.4.4 Review of Electronic Medical Record (EMR) and Medication Reconciliation

The EMR will be reviewed to check end organ function at time of study visits (i.e., recent bloods for liver function tests, urea, electrolytes and creatinine and full blood count) to confirm ongoing eligibility and to complete a medication reconciliation.

Medication reconciliation will include details of regular medications, dose, administration method and frequency. These will be entered into the *REDCap* database. Of interest we will document if patients have ingested drugs known to potentially affect hepatic drug metabolism. Pharmacists will conduct a thorough check using resources recommended by Medicines Information Centres (i.e., Flockharts drug interactions table - https://drug-interactions.medicine.iu.edu/MainTable.aspx).

**Timing of Assessment:** t_2_ at diagnosis, t_3_ at 2 months, t_4_ at 4 months, t_5/6_ at a maximum of two febrile episodes within 4-month period and at t_7_ at study conclusion (when finished treatment).

**Administration of Testing:** Performed by Academic pharmacist.

##### 11.4.5 Acceptability Survey

We will apply the (TFA) to (i) explore reasons for decline in consent for participating in pharmacogenomic and phenoconversion testing; and (ii) explore associations between decliners’ perceptions of acceptability and non-participation. The survey is provided in Supplemental Files 3-4.

**Eligibility**: Participant enrolled in PEGASUS.

**Timing of Assessment:** This is performed at t_7_ at study conclusion (when finished treatment).

**Duration:** 10 minutes

**Administration:** Survey will be disseminated via email through the *REDCap* database. The Academic pharmacist will ensure release date at the end of cancer treatment of the survey.

#### 114.6 Structured Questions for Decline in Consent

As described in 8.4.6 we will provide a short 5-minute survey for patients (Appendix 3) or parents/guardians (Appendix 4) at the time of decline of consent. The survey is provided in Supplemental Files 5-6.

**Eligibility**: Parent or carer of child who declined consent in PEGASUS.

**Timing:** At time of decline in consent

**Duration:** Less than 5 minutes

**Administration**: Structed interview; questions asked by study team at time of decline in consent.

If participants/guardians decline initial consent, verbal consent will be taken to answer the short decline consent survey. The study team will explain that a short decline to consent survey is not mandatory but in itself can help us to understand why the study is not acceptable to participants.

No participants/guardians details will be recorded as part of the decline in consent study. They will be given a study identifier be marked as “declined to consent” and only the answers to the questions will be recorded in REDCap.

### 12.0 RETENTION

Retention in the study is aided by the academic pharmacist/PhD student conducting weekly quality assurance assessments on data collection. Patients are contacted to meet any outstanding endpoints [i.e. TFA]. All study participants are provided with the ability to discuss their pharmacogenomic result at the time they are available with a member of the study team. Weekly newsletters are provided via the hospital electronic mail system for awareness of the broader enrolling health care professional team.

### 13.0 DATA MANAGEMENT

#### 13.1 Data Forms and Data Entry

PEGASUS Study results will be kept on a *REDCap* database. This is a secure database run by the Murdoch Children’s Research Institute. All paper files and consents will be kept locked away securely in a filing cabinet at each participating site.

Data will be collected in an identifiable manner. Upon entry of this data into *REDCap*, participants will be allocated a unique patient identifier. Consents and PGx reports will be uploaded to this database, in addition to key endpoint data collected as part of the study. Information extracted from the database will be de-identified however delegated members of the study team will have access to re-identify the study data for data entry and integrity purposes.

All database information is protected within Murdoch Children’s Research Institute using the *REDCap* database by a triple encryption process between the remote user, the server and the *REDCap* database. No data can leave the database in an identifiable format. All data extraction will be de-identified for all users at all times, thus privacy and confidentiality will be maintained. The database itself is password protected and only researchers involved in the study will have the ability to access information collected.

We are required to keep information collected as part of this study for at least fifteen (15) years following the last publication of the study. Following that time, all information that was collected will be securely destroyed.

#### 13.2 Governance for Data Management and reporting

The Principal Investigator (PI) is responsible for the design and conduct of the trial, preparation of protocol and revisions, preparation of case report forms (CRF) within REDCap, organising steering committee meetings, managing the clinical trials office, publishing study reports and membership of the trial management committee (TMC). The Steering Committee (SC) agrees on the final protocol, includes lead investigators from the two with responsibilities of recruitment of patients and liaising with the PI, reviewing study progress and if necessary, agreeing to changes of the protocol. TMC consists of the Principal Investigator and co-Investigator (MA) Academic Pharmacists (CM, EW, RD, DK) and PhD student. The team contribute to organisation of steering committee meetings, provide annual risk report to Human Research Ethics Committee (HREC) and Melbourne Clinical Trials Committee (MCTC), assess adverse events and serious unexpected adverse events (SUSAR) with appropriate reporting to HREC and MCTC. They are responsible for maintenance of the database, budget administration, HREC updates and amendments and randomisation.

#### 13.3 Data Endpoints and Databank Guidelines

Data endpoints to be recorded in the PEGASUS study database are described in Supplemental File 7 with databank guidelines provided in Supplemental File 8.

#### 13.5 Data Quality Assurance

An audit trail of transactions, available through the *REDCap database,* will be maintained to ensure data integrity and regulatory compliance. During data entry, each value entered will be subjected to a variety of quality control checks. Regular notifications identifying data quality issues and required follow-ups are regularly distributed by email to the national coordinator and then will be conveyed to the PI of the study. These data audits are discussed at weekly meetings with the pharmacogenomics study team.

### 14 STATISTICS METHODS

#### 14.1 Sample Size Estimation

As the primary outcome is feasibility and acceptability a sample size estimate is not required, however it should be noted that the investigators estimate that the recruitment period will be either adequate to permit an assessment of feasibility and acceptability, or to determine that a trial is not feasible due to low enrolment.

#### 14.2 Population to be analysed

The population analysed is Paediatric or AYA patient (6-25 years) with a new diagnosis of Lymphoma (Hodgkin or Non-Hodgkin subtype).

#### 14.3 Methods of analysis

In this single arm study group comparisons are not planned and therefore inferential statistics will not be undertaken. Descriptive statistics of the proportions as described in outcomes will be reported as percentages with binomial logistic confidence intervals derived via exact method. Counts and continuous variables will be described with measures of central tendency (median or mean depending on normality) and variance (interquartile range or standard deviation). Incidence of genotype phenotype mismatch during the study will be described as events/treatment course.

## METHODS: MONITORING

### 15 DATA MONITORING

#### 15.1 Trial Monitoring Group [TMG]

The PI is responsible for supervising any individual or party to whom they have delegated tasks at the study site. They must provide continuous supervision and documentation of their oversight. To meet this GCP requirement, a small group will be responsible for the day-to-day management of the study and will include at a minimum the PI, Academic Pharmacist and Research Assistant.

#### 15.2 Pharmacogenomics Steering Committee (PSC)

The Pharmacogenomics Steering Committee meet weekly to review the pharmacogenomic data for patients enrolled across the range of studies for MCRI pharmacogenomics team. This group consists of the Study PI Academic Pharmacist, Clinical Pharmacologist (as required) and a Bioinformatician, Paediatric Oncologist and co-PIs on this project. The meeting will serve the purpose of reviewing:

- number of participants enrolled.
- Schedule of upcoming assessments and assignment of Study team to undertake assessment.
- Compliance with testing requirements.
- Assess adverse events arising during the study.
- Addressing Quality Audits from *REDCap* database.

#### 15.3 Quality Control and Quality Assurance

The Sponsor-Investigator will have responsibilities in relation to quality management. The Sponsor-Investigator has developed SOPs identify, evaluate quality assurance aspects of the study. The Sponsor-Investigator will also implement quality control (QC) procedures, which will include the data entry system and data QC checks. This will include internal quality management of study conduct, data and biological specimen collection, documentation and completion.

### 16.0 HARMS

An adverse event will be defined as any untoward medical occurrence in a subject without regard to the possibility of a causal relationship. For the purposes of this study AE will be those events occurring within 2 hours of (a) taking the probe drug dosing OR (b) having procedures performed (blood draws). Study safety monitoring will be coordinated by TMG chaired by the PI and the study research assistant. Reported adverse events, assessed and classified individually and in consultation, will be subject to clinical and scientific judgment as per the TMG. Full event definitions and reporting guidelines are detailed in Supplemental File 9.

### 17.0 AUDITING

Auditing is performed weekly. Auditing ensures compliance with % enrolment, release of pharmacogenomic report, % completion of surveys, % completion of all timepoints, admissions for febrile neutropenia and % completion of study endpoints. Reports of study progress are due yearly to the Ethics Committee of The Sydney Children’s Hospital. The auditing is performed by an independent research assistant, separate to principal and associate investigators.

### 18.0 ETHICS AND DISSEMINATION

#### 18.1 RESEARCH ETHICS APPROVAL

This protocol and the informed consent document and any subsequent amendments will be reviewed and approved by the Sydney Children’s Hospital Human Research Ethics committee (HREC) prior to commencing the study. A letter of protocol approval by HREC will be obtained prior to the commencement of the study, as well as approval for other study documents requiring HREC review. This study was first approved by HREC in November 2023 (Supplemental File 10).

A letter of authorisation will be obtained from the RGO prior to the commencement of the research at The Royal Children’s Hospital. Institutional governance authorisation for any subsequent HREC-approved amendments will be obtained prior to implementation.

#### 18.2 PROTOCOL AMENDMENTS

This study will be conducted in compliance with the current version of the protocol. Any change to the protocol document or patient information and consent form that affects the scientific intent, study design, participant safety, or may affect a participant’s willingness to continue participation in the study is considered an amendment, and therefore will be written and filed as an amendment to this protocol and/or informed consent form. All such amendments will be submitted to the HREC, for approval prior to being implemented.

#### 18.3 CONSENT

Consent will be undertaken using the approved participant or parent/guardian consent form (Supplemental File 11). Consent process for Patients and Parents/Guardians on study Patients and parents of patients will be informed of the study by the Study PI or Pharmacist assigned to this study (either at Peter MacCallum or The Royal Children’s Hospital) following a screening of potentially eligible patients from the electronic medical record and weekly multi-disciplinary meetings with treating teams. Patients and their parents will be approached during routine outpatient appointments or, on the ward, if currently an inpatient. Patients and their parents will have the study explained to them and any questions will be answered. A screening of eligibility will be carried out and following this the parent will be left with a copy of the patient information and consent form (PICF).

In accordance with the National Statement, this study will utilise an active consent process. This will involve presenting potential participants with all the information they need to make an informed decision about participation. When the eligible participant or parent/guardian is invited to participate, they will be provided with detailed information on the purpose of the study, the extent of their involvement, the risks associated and the option to always refuse participation at any time will be explained.

The eligible participant or parent/guardian will be informed that enrolment into the study is a voluntary process and that the study is looking to map liver enzyme activities over time using sub-therapeutic administration of dextromethorphan and omeprazole across a maximum of 7 timepoints over 4 months.

The approach to informed consent:

- Participants and guardian(s) of participants will be informed of the study with an initial screening during their inpatient stay, or outpatient appointment, and left with a copy of the participant/parent PICF. This will be performed by the academic pharmacist, study research assistant or PI.
- Prior to performing any trial-specific procedure (including screening procedures to determine eligibility), signed consent(s) form will be obtained for each participant.
- The process will be that the investigator or delegated member of the trial team will discuss the study with relevant family members (i.e., parent/legal guardian).
- Potential participants as well as enrolled participants will be able to contact the studies academic pharmacist or PI and have the ability to ask questions and request clarifications at any time prior to or during the study via email.
- Where the patient is < 12 years of age, the parent/guardian will be provided with the parent/guardian PICF. The Parent/guardian will be asked to read and sign the study PICF if they agree and are willing for their child to participate.
- Where the patient is >12 years of age the parent/guardian will be provided with the parent/guardian PICF, and the patient will be provided with the Adolescent/Young adult PICF. Both parents/guardians and patient will be asked to read and sign the respective PICFs if they agree and are willing to participate.

#### 18.4 TREATMENT DISCONTINUATION, PARTICIPANT WITHDRAWALS AND LOSSES TO FOLLOW-UP

Participants and or their parent/legal guardian can withdraw consent for trial participation at any time during the trial.

Participants or legal guardians of participants can withdraw consent at any time of the study. This can be done by notifying the study PI (RCH: and Peter Mac) or delegated member of the study team. This can be done via any means the participant, or their legal guardian feel most comfortable communicating the desire to withdraw from the study.

Participants may discontinue trial treatment for the following reasons.

Participant/legal guardian request to discontinue trial intervention for any reason including undisclosed reasons the investigator decides to discontinue a participant from the trial intervention if the participant.

- Experiences a serious or intolerable adverse event such that continued trial intervention would not be in the best interest of the participant.
- Develops during the trial period symptoms or conditions listed in the exclusion criteria.
- Requires early discontinuation for another reason.

#### 18.5 STUDY CLOSURE

A participant is considered to have completed the study if they have completed all phases of the study including the last visit or the last scheduled procedure shown in the schedule of assessments.

The end of the study is defined as completion of the last visit or procedure shown in the schedule of assessments by the last participant at all study sites. At this stage, the study PI will ensure that all HRECs and RGOs are informed.

##### 18.5.1 TEMPORY HALT OR EARLY TERMINATION OF TRIAL

If the trial is prematurely terminated or suspended, the PI will promptly inform trial participants, HREC and RGO, the funding and regulatory bodies, and provide the reason(s) for the termination or suspension. Circumstances that may warrant termination or suspension include, but are not limited to:

- Determination of an unexpected, significant, or unacceptable risk to participants that meets the definition of a Significant Safety Issue (SSI).
- Insufficient compliance to protocol requirements.
- Data that is not sufficiently complete and/or evaluable.
- Significant advantage confirmed by participation in the trial, such that the trial should close early.
- In the case of concerns about safety, protocol compliance or data quality, the trial may resume once the concerns have been addressed to the satisfaction of the sponsor, HREC, RGO, funding and/or regulatory bodies.

The investigator may also withdraw all trial participants from the intervention if the trial is terminated. For the safety of all participants ceasing trial treatment in this event the protocol specified safety evaluations should be undertaken to capture new safety events and to assess unresolved safety events. All scheduled follow ups of trial participants should also occur following treatment discontinuation where possible.

##### 18.5.2 LOSSES TO FOLLOW UP

A participant will be considered lost to follow up if they fail to return for 2 scheduled visits and is unable to be contacted by trial staff. the following actions must be taken if a participant fails to the return to the clinic for a required trial visit.

- The site study staff will attempt to contact the participant and reschedule the missed visit and counsel the participant on the importance of maintaining the assigned visit schedule and ascertain of the participant wishes to and or should continue in the trial.
- Before a participant is deemed lost to follow-up the investigator or trial team will make every effort to regain contact with the participant (where possible, 3 telephone calls and if necessary, a certified email to the participant’s last known email address recorded on *REDCap*. These contact attempts should be documented in the participants medical record or trial file.
- Should the participant continue to be unreachable they will be considered to have withdrawn from the trial with a primary reason of lost to follow-up.

##### 18.5.3 REPLACEMENT

Participants who have been assigned a trial intervention will not be replaced.

### 19.0 CONFIDENTIALITY

Participant confidentiality is strictly held in trust by the PI, research staff, and the sponsoring institution MCRI and their agents. This confidentiality is extended to cover clinical information relating to participating participants.

The trial protocol, documentation, data and all other information generated will be held in strict confidence. No information concerning the trial, or the data will be released to any unauthorised third party, without prior written approval of the sponsoring institution.

Authorised representatives of the sponsoring institution may inspect all documents and records required to be maintained by the Investigator, including but not limited to, medical records (office, clinic or hospital) and pharmacy records for the participants in this trial. The Royal Children’s Hospital and Peter MacCallum Cancer Institute will permit access to such records.

All evaluation forms, reports and other records that leave the site will be identified only by the Participant Identification Number (SID) to maintain participant confidentiality.

Clinical information will not be released without written permission of the participant, except as necessary for monitoring by HREC or regulatory agencies.

### 20.0 PATIENT AND PUBLIC INVOLVEMENT

Patients and public were first involved in the design of the research at the time of applying for the funding through the Medical Research Future Fund (MRFF). The protocol and grant application were developed with consumer participants from the Victorian Paediatric Cancer Consortium (VPCC) including consumer chief investigators on the grant application. Over the last 12 months we have had 4 separate meetings with the VPCC Patient Advisory Group to inform on pharmacogenomics study progress, attain feedback on project methodology and consent.

## Data Availability

This is a protocol paper - so does not include data.

## 21.0 DECLARATION OF INTEREST

There are no identified financial or other competing interests for any investigators participating in this trial.

## 22.0 ACCESS TO DATA

The PI and steering committee will oversee the intra-study sharing process, at both the interim analysis and at the end of the trial.

The scientist performing phenotyping analysis will be blinded to the genotyping results and complete the analysis of the secondary endpoint blinded.

## 23.0 DISSEMINATION POLICY

### 23.1 Trial Results

At the conclusion of the trial, participants will be emailed the trial results to provide an overview of the findings. MCRI holds the primary responsibility for publication of the results of the trial.

## AUTHOR STATEMENT

The principal investigator for PEGASUS is RC in collaboration with MA whose team contributed to the writing of the study. RC, BF, TS, AG, MA and AS were involved in initial drafting. AH designed the bioinformatic workflow and pharmacogenomics report for the study. AS and CK designed laboratory assays for phenotyping. CM, DK, EW, RD, JS, AS, CK have contributed to refinement and editing of the protocol. DE is laboratory head and has had oversight to the trial rollout and design. All authors contributed to the protocol development and/or edited and approved the final manuscript.

## FUNDING

PEGASUS is funded through a nationally competitive Medical Research Future Fund Grant (EPCDRI Paediatric Cancer MRF/2007620) as part of the Pharmacogenomics discovery aim. This funding source had no role in the design of this study and will not have any role during its execution, analyses, interpretation of the data, or decision to submit results.

## COMPETING INTEREST STATEMENT

The authors have no competing interests to declare.

## ACKNOWLEDGEMENTS

This study is endorsed by the Victorian Paediatric Cancer Consortium (VPCC). VPCC facilitated consumer participation in pharmacogenomics discovery and barriers and enablers to implementation. We thank all of those who have contributed to the study. RC is supported by the Kids Cancer Project, The Royal Children’s Hospital Foundation, Victorian Paediatric Cancer Consortium, The Medical Research Future Fund and holds a Murdoch Children’s Research Institute (MCRI) Clinician Scientist Fellowship. MA is supported by an NHMRC Investigator Grant (GNT1196211) and holds a Peter MacCallum Cancer Centre Research Leader Fellowship (SSRL2023). D.A.E is a member of the Novo Nordisk Foundation Center for Stem Cell Medicine which is supported by a Novo Nordisk Foundation grant number NNF21CC0073729 and is supported by National Health and Medical Research Council of Australia (D.A.E), Heart Foundation of Australia and The Medical Research Future Fund.

MCRI is supported by the Victorian Government’s Operational Infrastructure Support Program.

## Notes

### Competing Interest Statement

The authors have declared no competing interest.

